# A new sensitive and robust next-generation sequencing platform for HIV-1 drug resistance mutations testing

**DOI:** 10.1101/2021.07.13.21260248

**Authors:** Bin Yu, Changzhong Jin, Zixuan Ma, Ziwei Cai, Tingsen Li, Dan Wang, Wenwen Xiao, Yanghao Zheng, Wanpeng Yin, Nanping Wu, Miao Jiang

**Author notes:** Equal contribution (Bin Yu, Changzhong Jin). (Bin Yu). (Jin Changzhong). (Miao Jiang). (Nanping Wu).

## Abstract

Next-generation sequencing (NGS) is a trending new platform which allows cheap, quantitative, high-throughput, parallel sequencing for minority variants with frequencies less than 20% of the HIV-1 quasi-species. In clinical setting, these advantages are crucial for choosing antiretroviral drugs with low genetic barriers and will potentially benefit treatment outcomes.

In this investigation, we implemented the Boxin HIV-1 NGS platform for genotyping the drug-resistance-associated variants in PR/RT regions. Plasmids with known mutations were used to analyze the accuracy, reproducibility, and reliability of the Boxin NGS assay. Variant frequencies reported by Boxin NGS and the theoretical value were highly concordant. The Bland-Altman plot and the coefficient of variation (7%) suggested that the method has excellent reproducibility and reliability. Sanger sequencing confirmed the existence of these known variants with frequencies equal or above 20%.

78 blood samples were obtained from AIDS patients and underwent PR/RT region genotyping by Sanger sequencing and Boxin NGS. 33 additional drug resistance mutations were identified by Boxin NGS, 23/33 mutations were minority variants with frequencies below 20%.

15 blood samples obtained from AIDS patients underwent PR/RT region genotyping by Sanger sequencing, Boxin NGS, and Vela NGS. The Bland-Altman plot suggested that the variant frequencies detected by Boxin and Vela were highly concordant. Moreover, Boxin NGS assay detected five more minority variants with frequencies ranged from 1% to 20%. In a series of samples collected from 2016 to 2017, Boxin NGS reported a M184V mutation with a frequency of 4.92%, 3 months earlier than this mutation was firstly detected by Vela NGS and Sanger sequencing.

In conclusion, Boxin NGS had good accuracy, reproducibility, and reliability. Boxin NGS was highly concordant with Sanger sequencing and Vela NGS. In terms of genotyping HIV-1 variants in PR/RT regions, Boxin NGS was more cost-efficient and appeared to have increased sensitivity without compromising sequence accuracy.

## Introduction

HIV-1 drug resistance (HIVDR) is the primary challenge to administrate an effective antiretroviral therapy (ART)^1–4^. Error-prone and rapid viral replication result in an extremely diversified HIV-1 quasi-species^3,5–7^. Under selection pressure, gene mutation-induced low-frequency drug resistance can surpass the effect of ART, become the dominant species in the host, give rise to a secondary resistance, and lead to a worse prognosis or a virological failure^2,4–6,8–11^. Therefore, identifying low-frequency HIVDRs is crucial for administering a successful ART^2,12–15^.

Sanger sequencing was considered as the gold standard for detecting drug-resistance mutations (DRMs)^16^. However, it inability of detecting low abundance drug-resistance mutations (LADRMs) present in less than 20% of the viral quasi-species is considered as a major obstacle^1,4,5,9,16,17^.

Compared to Sanger sequencing, NGS guarantees a cheaper, sensitive, parallel, and high-throughput genome sequencing method^4,16,18^. NGS allows a lower limit of detection of 1%, enhances the sequencing resolution, and deconvolutes complex HIV-1 quasi-species^5,6,19^. The ability of NGS-based genotyping in providing numerical data allows it to report the relative abundance of the variants and predict DRMs^5,17,20,21^. Recent studies have demonstrated the importance of identifying patients who are prone to virological failure due to an increasing evidence of LADRMs inducing immune system suppression, drug resistance aggregation, and death^5,22–24^.

In this study, we implemented and evaluated the Boxin HIV-1 NGS platform (Liangxin Biotechnology Development LTD) in terms of its accuracy, reproducibility, and reliability. We also compared the performance of Sanger sequencing, the Vela Diagnostics Sentosa SQ HIV genotyping NGS assay, and the Boxin NGS HIV-1 genotyping assay in testing PR/RT sequences.

## Methods

### Ethical statement

This study was reviewed and waived by the Clinical Research Ethics Committee of the First Affiliated Hospital, College of Medicine, Zhejiang University.

### Evaluation of the Boxin NGS using plasmids

Accuracy, reproducibility, and reliability of the Boxin NGS assay were investigated by genotyping amino acid variants in plasmids with known mutations. The plasmids also underwent Sanger sequencing to verify the existence of the variants. Three different HIV-1 plasmids were obtained from Sangon Biotech LTD, Shanghai, China. The plasmids were diluted to 0.2ng/ μL, and the wild type plasmid(YB002) was mixed with the plasmid with a single variant (YB003) or the plasmid with 7 variants (YB012) as shown in table 1. Each sample was prepared examined in triplicates, and the procedure was repeated twice by two different lab technicians who worked independently. Each sample was amplified by MiniAmp PCR (ThermoFisher). Sanger sequencing was performed by a reference laboratory (BGI Genomics, Beijing, China).

**Table 1.**
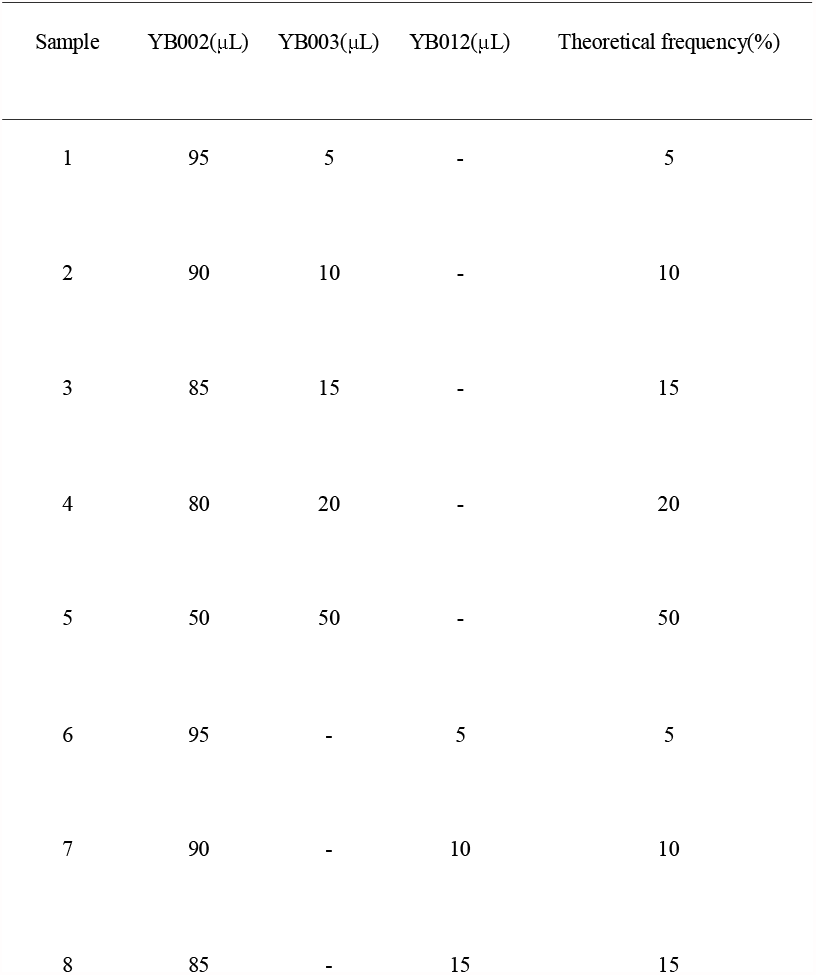
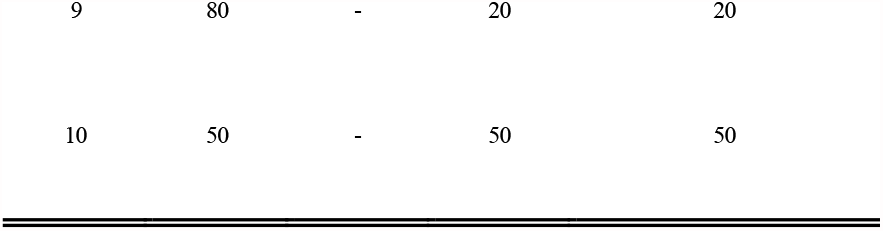
Constructing samples with known variant frequencies. YB002: wild type; YB003: including single amino acid variant, K65R; YB012: including seven amino acid variants, L100I, K101E, V106M, Y181F, Y188L, G190E, and M230L

### Patients and samples

87 blood samples were collected from AIDS patients treated at the First Affiliated Hospital, College of Medicine, Zhejiang University. All blood samples underwent Boxin NGS and Sanger sequencing. Another 15 samples were obtained from AIDS patients who were treated at the Beijing Ditan Hospital Capital Medical University, and underwent Sanger sequencing, Boxin NGS, and Vela NGS. Reverse transcriptase (RT) and protease (PR) sequences were genotyped. Protease inhibitors (PIs), nucleoside reverse transcriptase inhibitors (NRTIs), and nonnucleoside reverse transcriptase inhibitors (NNRTIs) associated DRMs were analyzed and reported by all three methods. Integrase inhibitors (INs) were not analyzed for the purpose of the study.

### Boxin NGS

Ion Plus Fragment Library Kit (Thermo Fisher Scientific, Waltham, USA) and Ion Shear™ Plus Reagents Kit (Thermo Fisher Scientific) were the reagents employed for library construction and DNA shearing, respectively.

RNA was extracted from 500 μL of each blood sample. Ion Torrent’s instruments (Thermo Fisher Scientific) were employed for deep sequencing. Two wild-type HIV-1 B subtype plasmids (positive and negative) were added to each run and was run in parallel with other samples as controls. 96 samples were examined for each run, including a positive and a negative control. RT-PCR was conducted to amplify the sample using the MiniAmp PCR Instrument (ThermoFisher). Library was by Ion Chef Instrument (ThermoFisher). A quantitative PCR was carried out to quantify the library. The bioinformatics pipeline was developed by ThermoFisher Scientific with optimization.

### Sanger sequencing and Vela NGS

500 μl of each blood sample was sent away to reference laboratories. Vela NGS assay was performed as per manufacturer’s instructions at Beijing Ditan Hospital Capital Medical University. Sanger sequencing was performed at a reference library (Tsingke Biotechnology Co., Ltd).

### Database and data analysis

Results were analyzed and reported using the NGS database developed by Liangxin (Beijing) Biotechnology Development LTD. The NGS database was constructed and optimized based on populated Chinese AIDS patients, which allowed identification and analysis of 256 PI, NRTI, NNRTI, and IN associated DRMs. Figures and tables were generated by Microsoft Excels.

## Results

### Evaluation of the Boxin NGS assay using plasmids

Regression analysis was applied to determine whether the variant frequencies reported by Boxin NGS and the theoretical values were concordant. The theoretical values of variant frequencies were graphed against the mean experimental results. The experimental data distributed close to the predicted values. The slope of the line was 0.8948, and the y-intercept was -0.7312. The coefficient of determination R^2^ was 0.9932 *(Fig.1A)*.

The reproducibility and reliability of the Boxin NGS assay were analyzed by comparing the mutation frequencies detected and reported by two lab technicians. Each plasmid sample was made in triplicates and the experiment was repeated twice by two examiners. The average value of the experimental variant frequency of each sample was reported by each examiner. The reproducibility was visualized by the Bland-Altman plot *(Fig 1B)*; the average variant frequency of the same sample was graphed against the difference between the variant frequencies reported by two examiners of the same sample. All data points distributed within the limits of agreement and closely to the line of bias. The line of bias was the average of the difference, which was -0.908. The lines of upper and lower limit of agreement were calculated as bias +/- 1.96*STDEV, which was found to be 2.504 and -4.321, respectively.

**Figure 1.**
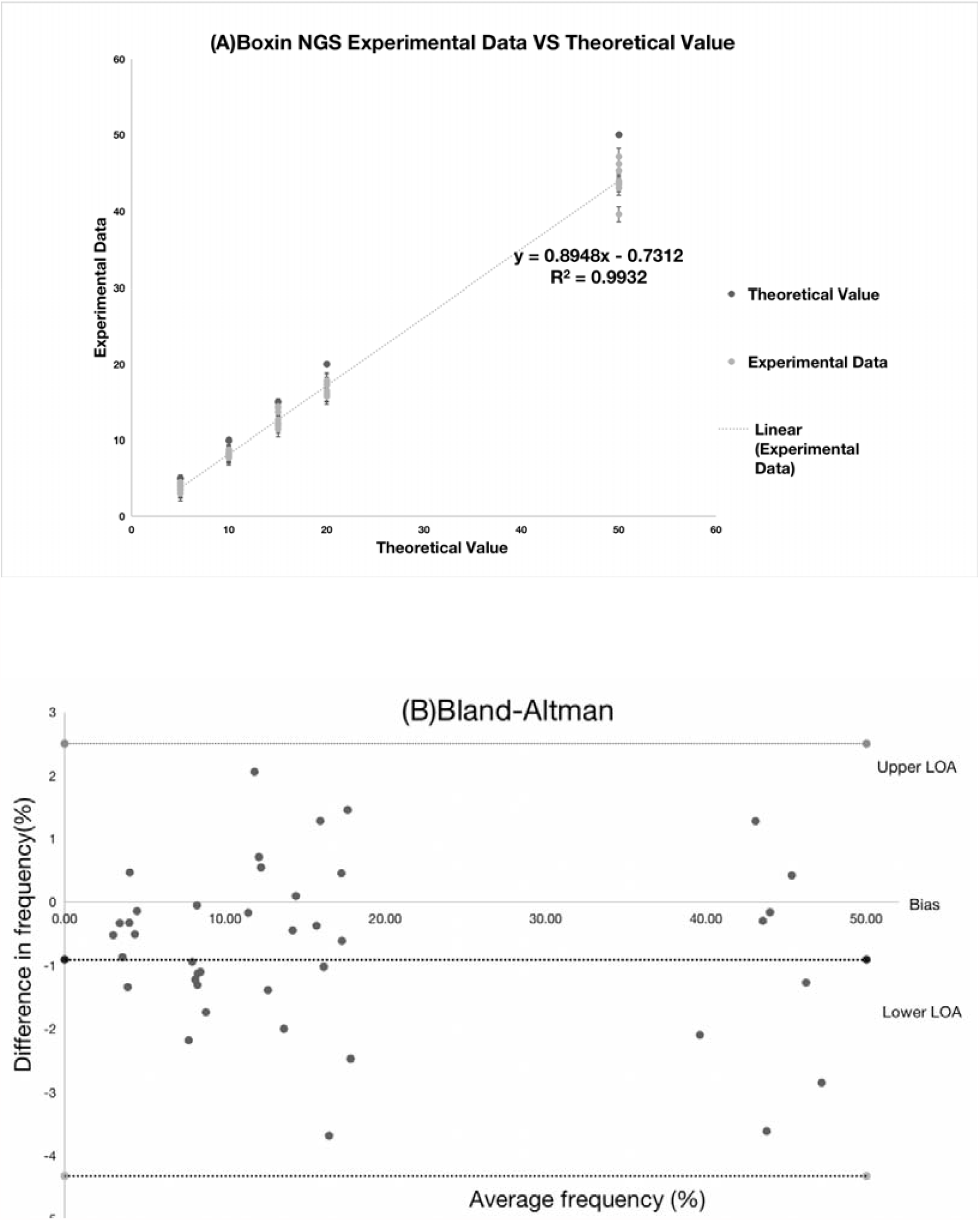
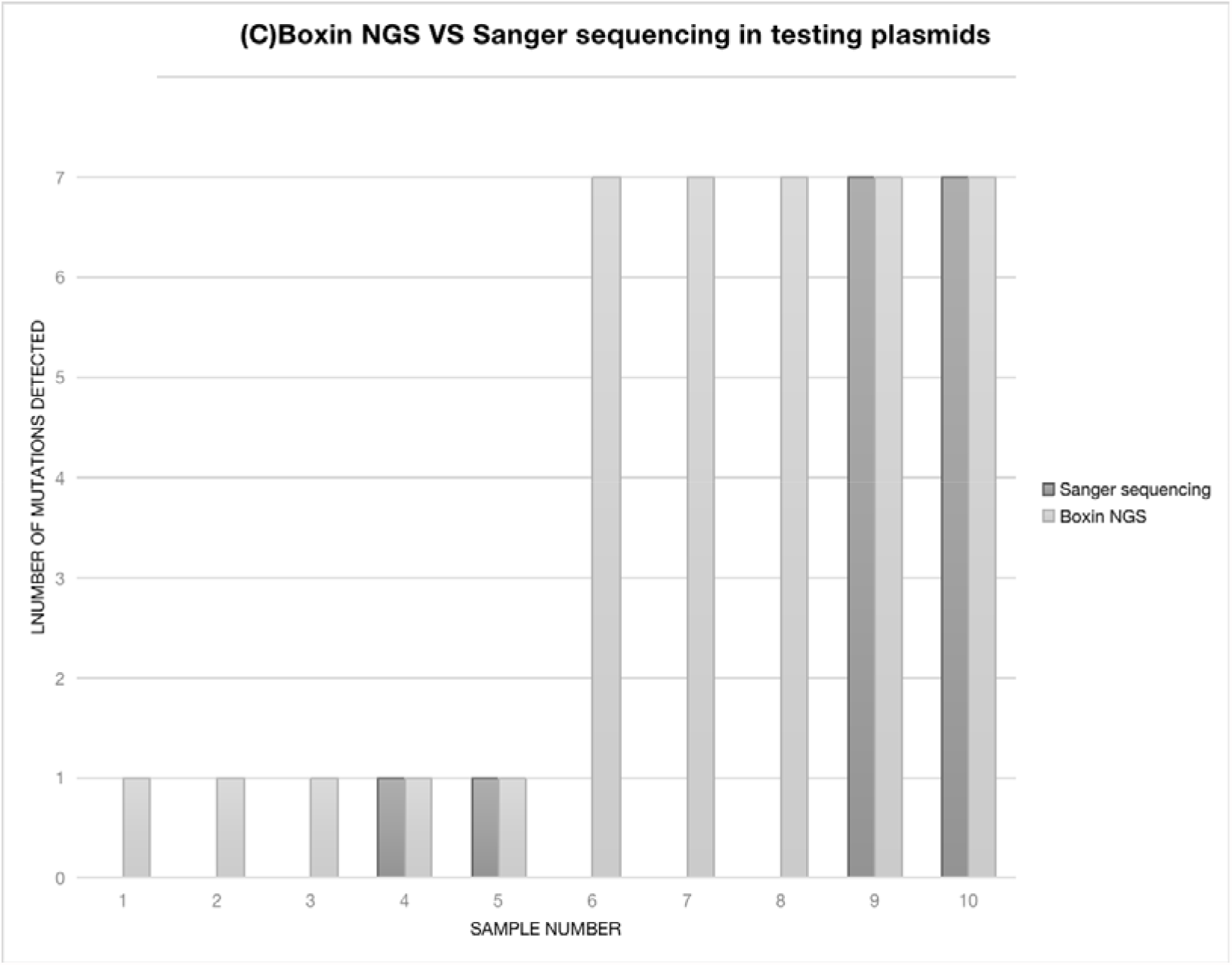
(A) Experimental data VS theoretical predictions of the Boxin NGS assay. The theoretical values were represented on the x-axis (theoretical mutation frequencies 5%, 10%, 15%, 20%, and 50%). The experimental results of the mutation frequencies with standard errors were labelled on the y-axis. (B) The Bland-Altman plot. Comparison of the Boxin NGS assay carried out by two examiners. The y-axis represented the frequency difference for a particular variant detected by two examiners. The x-axis represented the average frequency for a particular variant detected by two examiners. The x-axis represented the average frequency for a particular variant detected by two examiners.

The reliability of repeated measurements could be determined by the coefficient of variation. The coefficient of variation was calculated as the standard deviation divided by the mean variant frequencies reported by two examiners. The average coefficient of variation was found to be 7%.

Sanger sequencing assay was carried out to verify the mutations in the 10 samples constructed using plasmids. When the samples were prepared with variant frequencies equal and above 20%, Boxin NGS and Sanger sequencing reported the same mutations*(Fig. 1C)*. When the samples were prepared with variant frequencies below 20%, Sanger sequencing failed to detect any of the mutations. In comparison, the Boxin NGS assay was able to detect LADRMs with theoretical frequencies ranged from 5% to 20%.

### Comparison of Boxin NGS with Sanger sequencing using blood samples

Reverse transcriptase (RT) and protease (PR) sequences were genotyped by Sanger sequencing and Boxin NGS. 60/78 samples were identified to have NRTI or NNRTI associated DRMs, and no protease inhibitor (PI) associated DRM was detected. In total, Sanger sequencing identified 109 mutations, and Boxin NGS identified 142 NRTI or NNRTI associated DRMs. Variants detected by Boxin NGS alone were listed in bold. Boxin NGS detected additional 23/33 LADRMs with frequencies below 20%. 10/33 mutations had frequencies higher than 20%; 2/10 mutations had frequencies greater than 50% (table.2). Same amino acid variants caused by different changes of nucleic acids detected by Boxin NGS were underlined.

### Comparison of Boxin NGS with Vela NGS using blood samples

15 blood samples were sequenced by all three methods. RT/PR sequences underwent sanger sequencing and deep-sequencing, DRMs associated with PI, NRTI, and NNRTI drugs were identified. Corresponding variants frequencies were given by both Vela and Boxin NGS assays.

Overall, a total number of 20 mutations were identified by either or both NGS platform(s). Sanger sequencing only detected 5/20 mutations with frequencies higher than 20%. Boxin NGS detected 18/20 variants and Vela NGS detected 13/20 variants. 2 LADRMs with frequencies below 10% were detected by Vela NGS alone. 7 LADRMs with frequencies below 10% were reported by Boxin NGS alone(table 3).

**Table 2.**
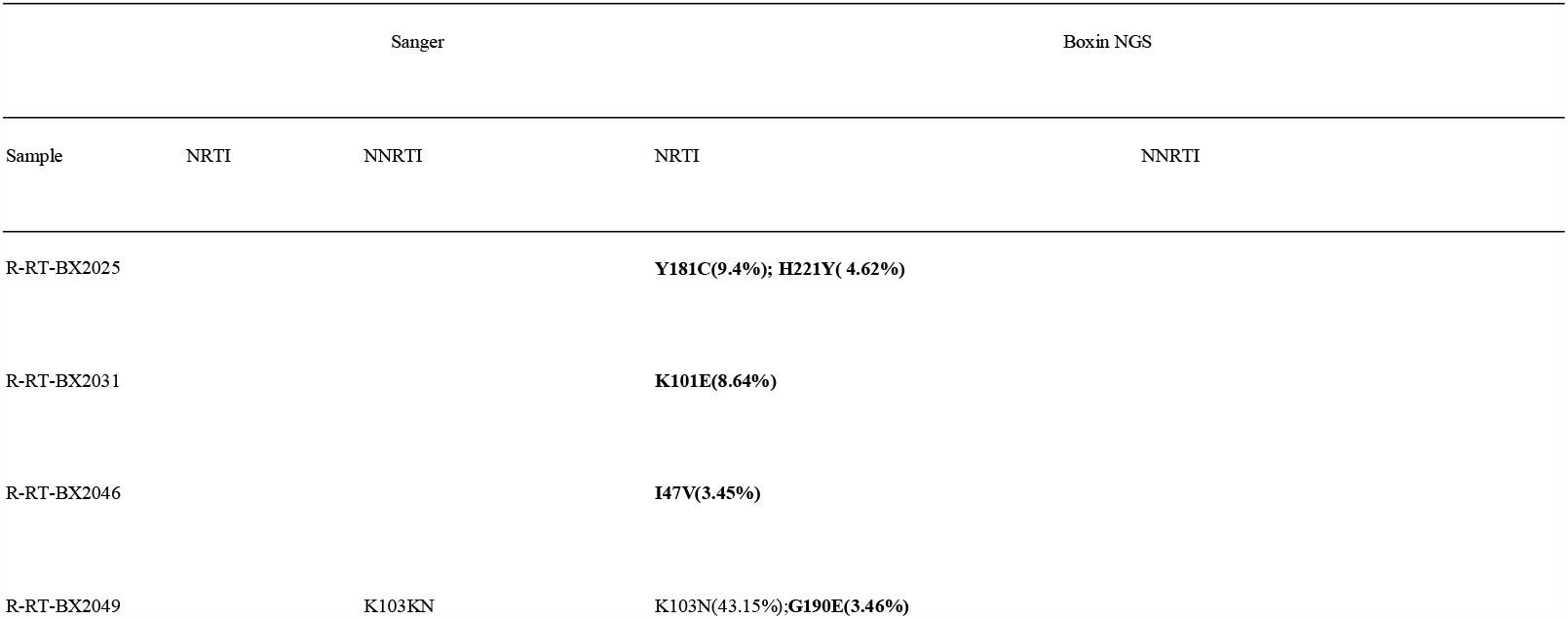
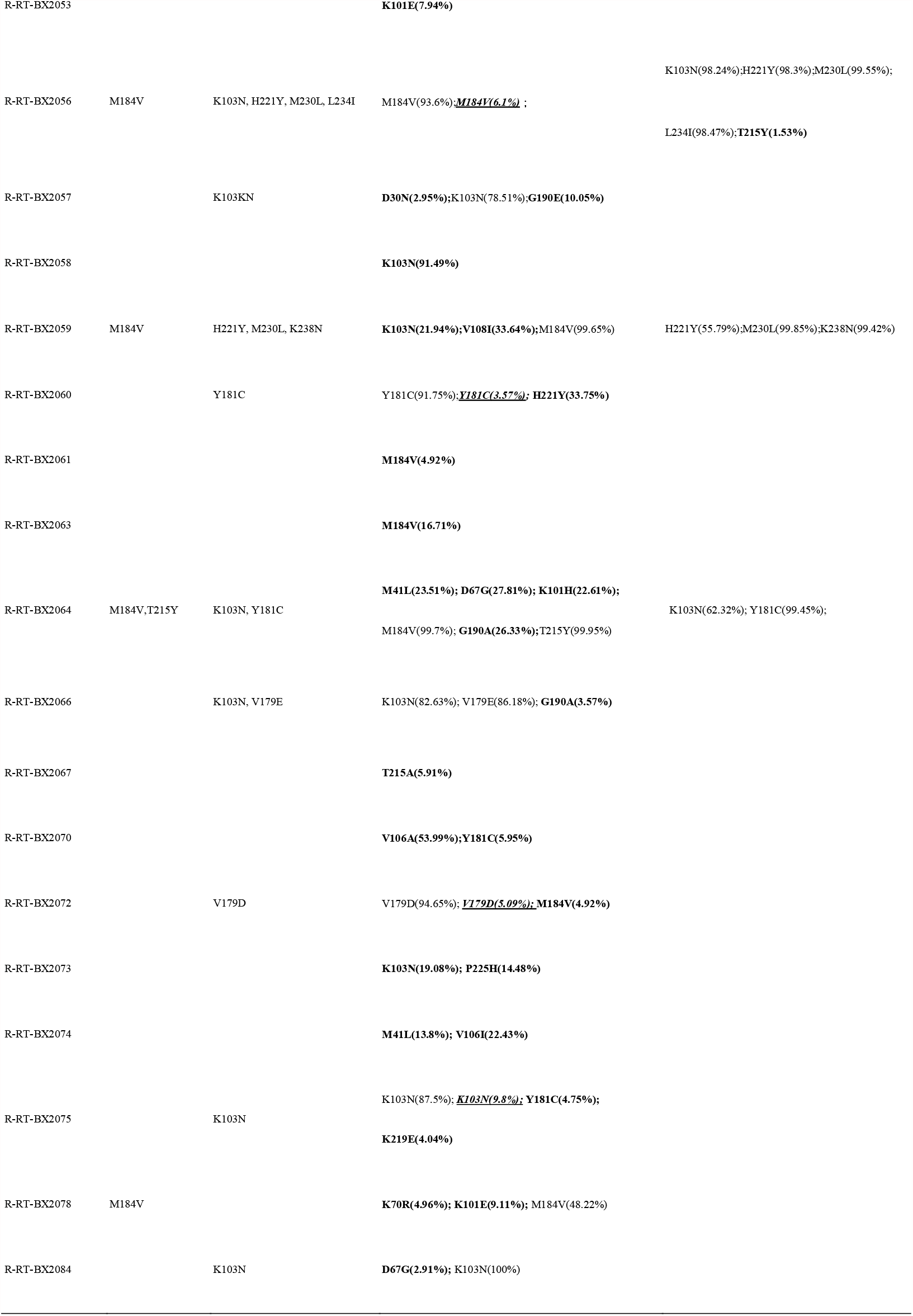
Disconcordant results between Sanger sequencing and the Boxin NGS assay in terms of NRTI and NNTRI associated DRMs. No PI DRM was identified by either of the two methods. Variants which were detected by Boxin NGS assay only were highlighted in bold. Same amino acid variants detected by Boxin NGS but caused by different mutated nucleic acids were underlined.

**Table 3.** Number of mutations detected by Boxin and Vela NGS platform.

| Mutation Frequency | Boxin | Vela |
| --- | --- | --- |
| 1%-5% | 5 | 2 |
| 5%-10% | 3 | 2 |
| 10%-20% | 2 | 0 |
| >20% | 8 | 9 |
| Total | 18 | 13 |

Boxin NGS and Vela NGS detected 11 common mutations, and the numerical values were analyzed statistically. A Bland-Altman plot was graphed to demonstrate the agreement of the numerical data between the two methods *(Fig.2)*. The lower limit of agreement was -0.252, the upper limit of agreement was 0.213, the bias was -0.0194. All points distributed close to the line of bias, and no point exceeded the limit of agreement.

**Figure 2.**
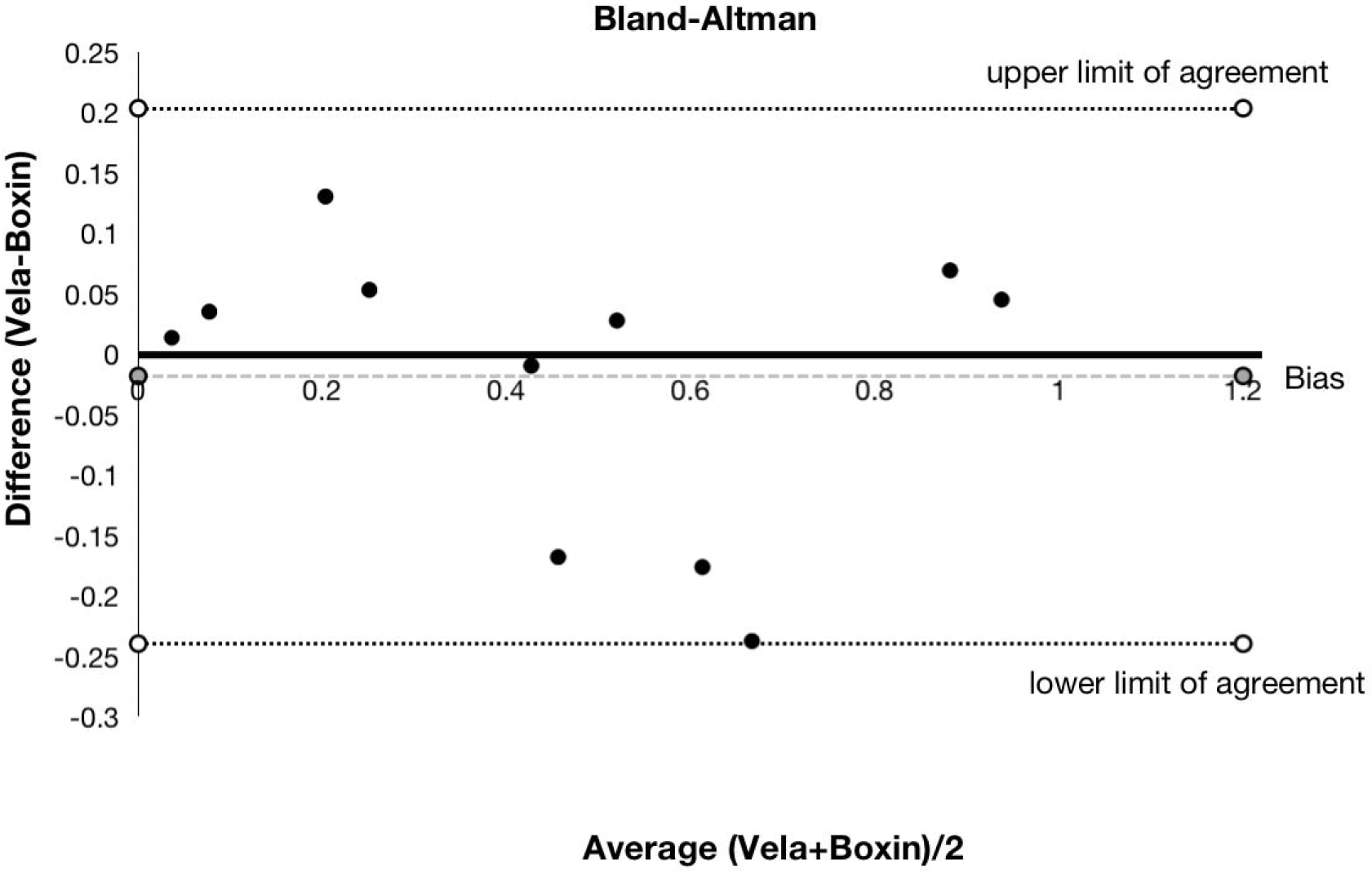
The Bland-Altman plot. Comparison of Boxin and Vela NGS methods in quantifying drug-resistance associated mutations. The y-axis represented the variant frequency difference detected by Boxin and Vela NGS. The x-axis represented the average variant frequency detected by Boxin and Vela NGS.

Four blood samples were collected from a patient at different periods between 2016 and 2017(table 4). In the second quarter of 2016, no variant was detected or reported by neither sequencing method.

**Table 4.** Mutations detected in blood samples collected from a single patient in different periods between 2016 and 2017.

| HIV-1 infected case 3 | SS | Vela NGS | Boxin NGS |
| --- | --- | --- | --- |
| 2016 2nd Quarter | None | None | None |
| 2016 4th Quarter | None | None | **M184V(4.92%)** |
| 2017 1st Quarter | M184MV | M184V(53.39%) | M184V(50.58%) |
| 2017 3rd Quarter | M184V | M184V(99.01%) | M184V(99.45%) |

However, at the end of year 2016, Boxin NGS detected a minority variant M184V with a frequency of 4.92%, while Sanger sequencing and Vela NGS failed to detect this low-frequency variant. Boxin NGS detected this LADRM 3 months prior to other sequencing platforms. In the 1^st^ quarter of 2017, Sanger sequencing reported a non-specific result as M184MV, and the Vela assay reported the mutant M184V with a frequency of 53.59%. Half a year later, the variant frequency reached 99%.

## Discussion

In this study, we evaluated the Boxin NGS assay, and compared the performance of the Boxin NGS assay with the conventional Sanger sequencing method and the FDA-approved Vela NGS assay.

To evaluate the accuracy, reproducibility, and reliability of the Boxin NGS assay, 10 plasmid samples were made with different variant frequencies. Samples were made and examined in triplicates, and the experiment was repeated by two examiners who worked independently. The theoretical values were graphed against the experimental results as illustrated in figure 1A. The slope of the linear regression line was 0.8948, close to 1; the observed values also distributed close to predicted values, suggesting they were highly concordant. The coefficient of determination R^2^ was 0.9932, suggesting variables had an excellent correlation. Reproducibility was defined as the variation of the same measurement under different conditions^25^. The reproducibility of the Boxin NGS assay was visualized by comparing the two sets of measurements carried out by two examiners using the Bland-Altman plot^25,26^. All data points distributed closely to the line of bias, suggesting the assay had an excellent reproducibility *(Fig.1B)*. The degree of inaccuracy between repeated measurements could be expressed by the coefficient of variation. The average coefficient of variation was calculated to be 7%, suggests that the results had an excellent reliability and accuracy^25^.

60/78 blood samples underwent Sanger sequencing and Boxin NGS were identified to have NRTI or NNRTI associated DRMs, and no protease inhibitor (PI) associated DRM was detected. Compared to Sanger sequencing, Boxin NGS detected additional 33 DRMs, 23/33 mutations with frequencies blow 20%. Sanger sequencing also failed in detecting two mutations with frequencies above 50%. Sanger sequencing also gave non-specific results such as Y188YL, M184MV. This could happen when peaks share a similar intensity. Boxin NGS also reported the frequency of the same amino acid variant caused by different changes of nucleic acids as shown in table 2. Compared to Sanger sequencing, Boxin NGS was able to specific and numerical results, and the platform was more sensitive in detecting LADRMs.

15 random blood samples collected from different AIDS patients underwent all three sequencing methods. Compared to Sanger sequencing, Vela NGS and Boxin NGS were sensitive enough to detect and quantify LADRMs with frequencies below 20%. The Bland-Altman plot was widely used to illustrate the agreement between two instruments or two methods^9,25–28^, and was applied to determine the degree of agreement of Boxin NGS and Vela NGS*(Fig.2)*. The average value of the variant frequencies reported by both platforms were plotted against the difference between the same measurements. The scattering points distributed close to the line of bias without exceeding the lines of limit of agreement, implying that the two methods were highly concordant^25,29–31^. 5 additional LADRMs with frequencies at or below 10% were detected by Boxin NGS, suggesting that Boxin NGS platform performed better at detecting LADRMs. As shown in table 4, Boxin NGS detected a low-frequency M184V variant with a frequency of 4.92%, and other methods failed to detect this variant. The result was later confirmed by Sanger sequencing and Vela NGS, three months after the variant was first reported by Boxin NGS. The mutation kept developing and became the dominant mutation of the HIV-1 quasi-species; the frequency reached to 99% in 9 months. Early detection of LADRMs can significantly reduce the possibility of virological failure, and potentially benefit the treatment outcome. This case highlighted the reliability and sensitivity of Boxin NGS in detecting LADRMs. Boxin NGS method was also more cost-efficient. The cost per sample for Sanger sequencing ranged from £430 up to £1050^32^. Vela NGS costed $400 for each sample^5^, while Boxin NGS only costed $40 per sample, which was 90% less than Vela NGS.

In conclusion, the new cost-efficient, robust Boxin NGS appeared to be more sensitive in reporting genomic variants, especially for LADRMs, without compromising the accuracy. The ability of early identifying and predicting DRMs of Boxin NGS allowed it to have more potentials in improving ART outcomes and preventing treatment failures. Although the HIV-1 LADRMs was confirmed to have correlations with virologic failure^10^, more cohort studies should be conducted to clarify the threshold for switching treatment and how to respond to the presence of LADRMs.

## Data Availability

The datasets used and/or analyzed during the current study available from the corresponding author on reasonable request.

## Acknowledgments

We thank Beijing Ditan Hospital Capital Medical University for providing Vela NGS results. We appreciate their hard work and their diligence to patient care.

## Author Contributions

YB, MZ, JC, ZY, YW, and WN designed the study. YB, MZ, JC, ZY, YW, WN, LT, WD, and XW performed molecular and genotyping experiments. YB, MZ, LT, JM, WD, and XW provided the logistics in implementing the Boxin NGS assay. CZ, MZ, and LT analyzed the data. CZ wrote and drafted the manuscript. All authors read and validated the manuscript.

